# Remission and clinical patterns of systemic lupus erythematosus (SLE) in Pakistan: a retrospective cohort study

**DOI:** 10.1101/2022.11.28.22282863

**Authors:** Mohammad Saeed, Aneela Pasha, Syed Hussain Azhar Rizvi, Maham Munawar, Zehra Abaad Elias, Muhammad Irfan Shafi, Tariq Gazdar, Maryam Ali Lakhdir

**Author notes:** **Correspondence:** Dr. Mohammad Saeed, M.D., Consultant Rheumatology and Immunogenetics, ImmunoCure Center for Inflammatory Diseases, Suite # 102, 1st Floor, Building 24-C, Lane 1, Ittehad Commercial Area, DHA Phase 6, Karachi, Pakistan, Web:www.immunocure.pk, 0331-8822712.

## Abstract

**Objectives:** Primary objective was to investigate clinical features and biomarkers associated with severe systemic lupus erythematosus (SLE). The secondary objective was to identify patterns of SLE remission. ’

**Methods:** A retrospective study of 200 SLE patients (2014–20) from ImmunoCure Center was conducted. Patients fulfilled ACR criteria 1997 for SLE classification. SLEDAI-2K categories mild-moderate (score <=10) and severe (score >10) were used as outcome for the primary objective. Predictors of severe SLE were evaluated by multivariate logistic regression analysis. For the secondary objective, we evaluated 94 records with follow-up time >1year. Remission status (Yes/No) was based on DORIS criteria. Survival regression was performed using Kaplan Meier curve.

**Results:** Significant predictors of severe SLE were male gender (OR 4.1; 95% CI: 1.2, 13.5), oral ulcers (OR 6.9; 95% CI: 2.8, 17.1), alopecia (OR 2.1; 95% CI 1.0-4.1), nephritis (OR 4.5; 95% CI: 1.9-11.4), ESR >30mm/hour (OR 2.3; 95% CI: 1.2-4.4) and aCL antibodies (OR 2.4, 95% CI 1.0 -5.9). The mean duration of follow-up was 41±19 months. Remission on treatment was achieved in 66% of 94 patients, while off treatment in 21% with a mean post-remission follow-up of 18±15 months. For every one-month increase in the duration of follow-up, the hazard of time to remission increased by 4% (95% CI 0.95-0.98; *P*<0.001). Factor analysis identified 4 SLE subtypes.

**Conclusion:** A clinical model including aCL antibodies is presented here that predicts severe SLE. Remission is possible even in severe SLE in LMIC with adequate immunosuppression and persistent follow-up.

## Introduction

Systemic lupus erythematosus (SLE) is a chronic autoimmune disease involving multiple organ systems. The disease is characterized by autoantibody presence and immune complex deposition (1). With multi-organ involvement, SLE presents as a clinically heterogeneous and unpredictable disease, ranging from mild rash to severe vasculitis lesions, arthritis, brain lesions, and glomerulonephritis (2). The clinical heterogeneity can often cause delays in diagnosis and subsequent damage accrual. Patients with SLE are at increased risk of developing complications, including cardiovascular and renal disease (3). The fully adjusted all-cause mortality is 1.85 times in SLE patients as compared to the general population (4).

The global estimates for SLE incidence vary considerably, from 0.3 per 100,000 per year in Ukraine to 517.5 per 100,000 among Afro-Caribbean people living in the UK (5-8). A study on the epidemiology of SLE in Asia found high incidence in Pakistan, Iran and China (30-50 per 100,000), whereas India, Japan and Saudi Arabia showed a markedly lower prevalence of 3.2-19.3 (9). Worldwide prevalence estimates were found to be 9 times higher among females than among males (10). It is debated whether SLE is more common or more severe in the Asian population as compared to Caucasians, primarily due to methodological heterogeneity, as well as genetic, environmental and other confounders that may affect disease prevalence and severity (9, 11, 12).

Given the complex heterogeneity of SLE, several studies have attempted to classify the disease into subgroups for purposes of improved clinical management. These subgroups have been based on clinical patterns (13, 14), auto-antibody profiling (13-15), organ domains (16), flares (17, 18), remission patterns (19), and treatment response (20). Using unsupervised cluster analysis, Diaz-Gallo et al (13) identified four distinct subgroups based on auto-antibody profiling. Interestingly, these subgroups also had differential clinical patterns indicating that SLE may exist as distinct sub-phenotypes.

Medical management of SLE involves antimalarials, oral corticosteroids, immunosuppressive agents and monoclonal antibodies, as defined by the ACR/EULAR SLE treatment guidelines (2). Treatment patterns are not remarkably different in Asia/ Pakistan from those in other countries (12, 21). The goals of treatment are complete remission based on SLE DORIS framework (22), with sustained improvement in quality of life and reduction in target organ damage. As prognosis and survival of SLE patients has improved significantly in the last decade, there is a shift in focus to remission and ‘Treat-to-Target (T2T) strategies for SLE patients, as with other chronic diseases (23). T2T strategies are not entirely drug-based, and include approaches, such as long term follow-up and social support (24). Our study aimed to characterize disease severity, clinical manifestations, treatment patterns and remission in a longitudinal cohort of patients in Pakistan, in order to define SLE in greater detail that is generalizable to a low-middle income country (LMIC).

## Methods

### Study Design

A retrospective cohort study was conducted to evaluate the clinical pattern and treatment response of SLE in an autoimmune disease referral center in Karachi. Records were analyzed from 2014 to 2020. Medical records of patients who met the American College of Rheumatology criteria 1997 for SLE classification (25) were included in the study. Records of SLE patients with overlap syndrome or incomplete records were removed from the analysis. The final sample size was 200.

### Study Setting

Karachi is the largest metropolitan city of Pakistan. It has a population of ∼20 million, which is served by over 20 major private, public and non-profit hospitals. Much of the staggering non-emergency patient load is cared for in private clinics scattered around the city. ImmunoCure is one of the few private autoimmune disease referral centers in Karachi. Established in 2014, ImmunoCure is known for its excellence in autoimmune health care and unique digitalization methods, including electronic medical records (EMR), telehealth and e-pharmacy services.

### Sampling Size and strategy

A total of 350 records at ImmunoCure met the ACR 1997 criteria for SLE between 2014 and 2020. Of these, 150 had overlap syndrome, including rheumatoid arthritis and Sjogren’s syndrome. Further, 106 patients did not follow beyond the first year or had incomplete records and were removed from the disease remission analysis. The final number of complete records analyzed for SLE presentation was 200, and for disease remission status was 94 (Figure 1). The patients were seen every 1 to 3 months (depending on disease severity), and in some cases additionally as clinically required. Missed appointments were noted in the EMR system. The same Rheumatologist also evaluated SLE disease activity based on SLEDAI score (26). The following laboratory parameters were evaluated as per standard clinical protocols: complete blood counts (CBC); erythrocyte sedimentation rate (ESR), C-Reactive Protein (CRP), liver transaminases, serum creatinine, urine dipstick, ANA (Hep2-immunofluorescence), ANA subsets (Immunoblot, Euroimmun) including dsDNA antibodies, anti-cardiolipin (aCL) IgG, IgM and SLEDAI score. Patients in clinical remission on hydroxychloroquine (HCQ) and low dose steroids only, who chose to go off all medications were allowed to do so after explaining the risks and with strong recommendation for follow-up. Open and detailed discussion on the disease and treatment were encouraged throughout.

**Figure 1:**
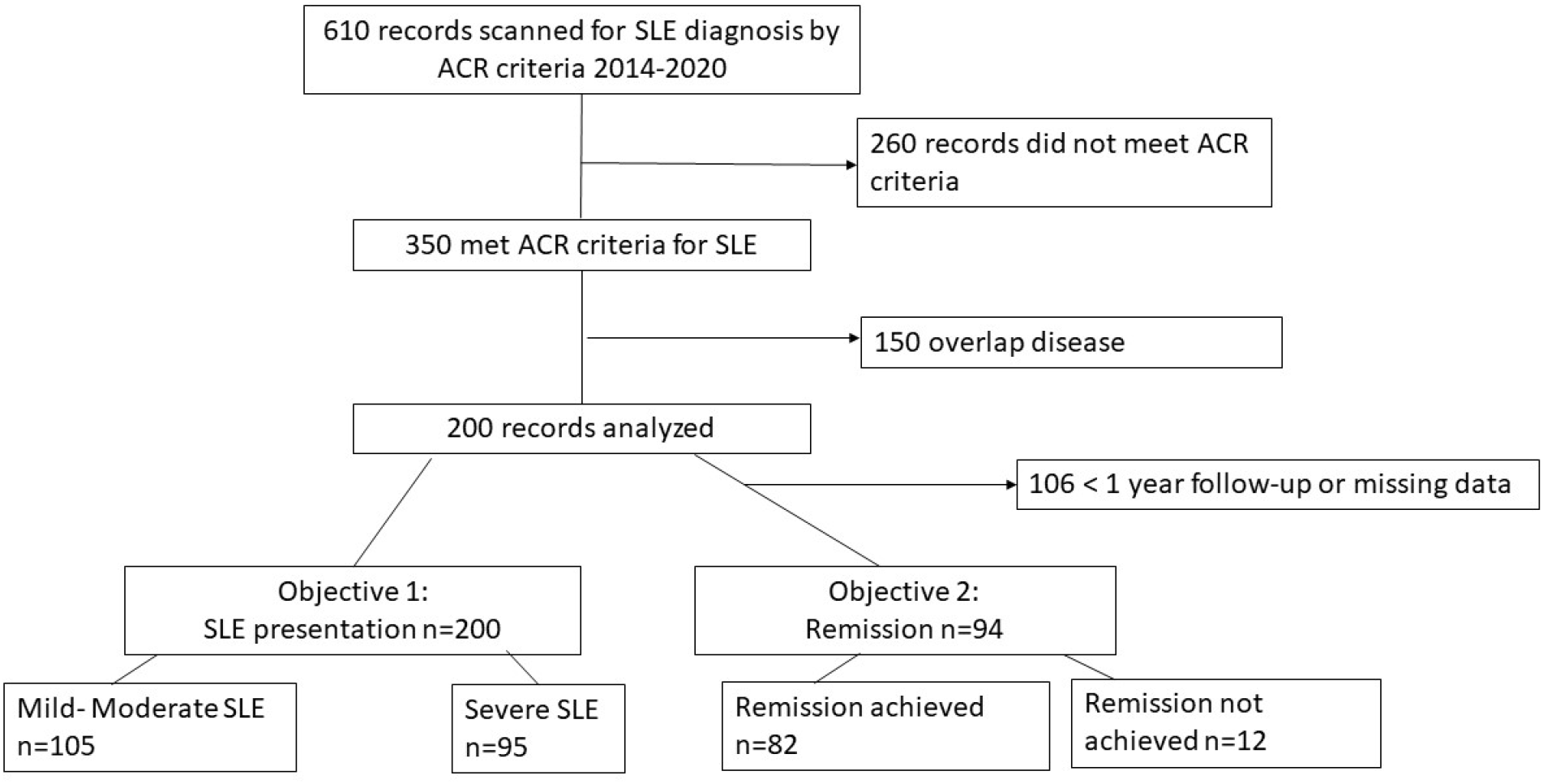
Flowchart of patient records analyzed in this retrospective cohort study.

### Data Collection and Management Methods

Data were collected in person by a dedicated research associate using a structured data retrieval form. The form was used to extract relevant data from the EMR. Each EMR at ImmunoCure Center has a unique identifier to guarantee complete confidentiality and privacy. The principal investigator (PI) of this study checked for completeness, inaccuracies, and illogical entries daily. The PI and another physician at ImmunoCure cross checked data entered in the data retrieval form with data from the EMR. This study was approved by the ImmunoCure ERC (IC-ERC-001-2022).

### Study Variables

The primary outcome was SLEDAI score that was categorized as mild (SLEDAI 1-6: coded 1), moderate (SLEDAI 6-11: coded 2) and severe (SLEDAI >11: coded 3), based on criteria defined previously (27). Mild and moderate categories were merged, and used as the reference category for severe SLE. SLEDAI was measured by SLEDAI-2K tool (Systemic Lupus Erythematosus Disease Activity Index) which contains 24 items, of which 16 are clinical and 8 are based solely on laboratory test results (26). It includes evaluation of specific manifestations in nine organ systems. SLEDAI has shown validity and reliability globally (28). SLEDAI has a Cronbach alpha between 81.7% and 100%, indicating the observer’s reliability of the scale is commendable (26). Independent variables included frequency of SLE features, ANA patterns, ANA subset antibodies, ESR (mm/hour) and CRP (mg/dL) levels. Subjects were classified into yes/no based on presence or absence of an antibody.

The secondary outcomes were remission (yes/ no) and time to remission (months). Remission was based on DORIS criteria (22). Remission on treatment (RONT) included immunosuppressives (IS) with Prednisolone (Pred) 5mg or 2.5mg and remission off treatment (ROFT) was defined as a patient being off all drugs (Pred 0 and no IS). Remission (yes) included RONT and ROFT categories. Additionally, total Pred and IS doses were calculated using software developed by the PI (29). Remission was evaluated only for patients with greater than one year follow-up. Remission status was noted by the same physician who cared for the patient. Time to remission was modeled against disease severity. Time to remission was defined as the time from diagnosis to DORIS remission.

### Statistical Analysis

Statistical analyses was performed using STATA version 17.0 software. Median and ranges were reported for continuous biomarkers (skewed data). Kruskal-Wallis test was used to compare the medians in each category in case of skewed distributions (ESR and CRP). The three continuous variables (age, CRP and ESR) were then transformed into categorical variables according to their reference values. CRP<0.5mg/dL; ESR<30 mm/hour; and age <40 years. Frequencies and percentages were used to describe categorical data. ANOVA was used to compare differences in age and Pearson’s chi square test was used to compare categorical explanatory variables across SLEDAI severity. Fisher’s Exact test was used to compare categorical explanatory variables when the expected cell count was less than 5 per category. The univariate and multivariate cumulative logit models were fitted with severe disease compared to mild-moderate as reference. Potential predictors of severity were investigated using univariate logistic regression first (using a cutoff of *P*<0.25). Furthermore, a forward stepwise multivariate logistic regression analysis (using a cutoff of *P*<0.05) excluding variables which were not significant in univariate cumulative logit model was conducted. Plausible interactions between significant variables were tested at 10% significance level. Hosmer–Lemeshow (HL) test was used to check the fit of the final model.

Cox Proportional Algorithm was used to evaluate the odds of remission by duration of follow-up. Kaplan Meir curve was generated for time to remission analysis and the log-rank test was applied to compare the remission status among different severity illness groups. The hazard ratio with 95% confidence interval (CI) was estimated. A two-sided α of less than 0.05 was considered statistically significant. Lastly, factor analysis was used to reduce data on SLE clinical features and antibodies and form meaningful disease subgroups for this population.

## Results

A total of 200 SLE records (2014 to 2020) from ImmunoCure Center in Karachi were analyzed. Of these 16.5% (n=33) had mild SLE disease activity, 36% (n=72) had moderate disease activity, and 47.5% (n=95) suffered from severe SLE. Mild and moderate SLE were merged into a single category and compared with severe SLE. Clinical data for these two SLE disease activity groups are presented in Table 1. The mean age of the patients was 37.5±13.8 years. There was no difference in age across SLE disease severity groups (*P*=0.4). Of the 16 males, 10 presented with severe disease. Mean duration of follow-up was 21.4±23.1 months.

**Table 1:** Clinical features and Biomarkers of SLE across SLEDAI severity categories.

| Predictors of disease severity | Total<br>(N=200) | Mild-Moderate SLE<br>(SLEDAI ≤10)<br>n=105 (52.5) | Severe SLE<br>(SLEDAI >10)<br>n=95 (47.5%) | P value |
| --- | --- | --- | --- | --- |
| Age (mean ± SD) | 37.5±13.8 | 38.3±12.9 | 36.7±12.9 | 0.4 |
| Female | 184 (92) | 99 (94.3) | 85 (89.5) | 0.2 |
| Male | 16 (8) | 6 (5.7) | 10 (10.5) |  |
| **Clinical features** (Freq % positive): |  |  |  |  |
| Arthritis | 169 (84.5) | 89 (84.8) | 80 (84.2) | 0.5 |
| Alopecia | 107 (53.5) | 51 (48.6) | 56 (58.9) | 0.1 |
| Oral Ulcers | 39 (19.5) | 8 (7.6) | 31 (32.6) | <0.001** |
| Rash | 111 (55.5) | 54 (51.4) | 57 (60.0) | 0.2 |
| Cytopenias | 89 (44.5) | 36 (34.3) | 53 (55.8) | 0.002** |
| Clots | 9 (4.5) | 3 (2.9) | 6 (6.3) | 0.2 |
| Neurological Disease | 35 (17.5) | 3 (2.9) | 32 (33.7) | <0.001** |
| Nephritis | 33 (16.5) | 9 (8.6) | 24 (25.3) | 0.001** |
| Interstitial Lung Disease | 20 (10) | 10 (9.5) | 10 (10.5) | 0.41 |
| Gastrointestinal System | 24 (12.0) | 8 (7.6) | 16 (16.8) | 0.04* |
| Follow-up Time (mean months ± SD) | 21.4±23.1 | 18.9±20.3 | 23.5±25.7 | 0.3 |
| **Biomarkers** (Freq % positive) |  |  |  |  |
| ESR>30 mm/hour Freq (% positive) | 102 | 42 (40.0) | 60 (63.2) | 0.001** |
| CRP >0.5 mg/dL Freq (% positive) | 121 | 54 (51.4) | 67 (70.5) | 0.006** |
| ANA (Freq % positive) |  |  |  |  |
| Homogenous | 103 (51.5) | 39 (37.1) | 54 (56.8) | 0.2 |
| Speckled | 24 (12) | 18 (17.1) | 6 (6.3) | 0.02* |
| ANA negative (Freq %) | 68 (34) | 33 (31.4) | 35 (36.8) | 0.8 |
| ANA Subsets (Freq % positive) |  |  |  |  |
| Mi2 | 23 (11.5) | 15 (14.3) | 8 (8.4) | 0.38 |
| KU | 35 (17.5) | 22 (21.0) | 13 (13.7) | 0.09 |
| RNP | 25 (12.5) | 11 (10.5) | 14 (14.7) | 0.63 |
| Sm | 29 (14.5) | 11(10.5) | 18 (18.9) | 0.22 |
| SSA | 48 (24) | 25 (23.8) | 23 (24.2) | 0.91 |
| Ro52 | 17 (8.5) | 10 (9.5) | 7 (7.4) | 0.86 |
| SSB | 19 (9.5) | 9 (8.6) | 10 (10.5) | 0.38 |
| Scl-70 | 15 (7.5) | 10 (9.5) | 5 (5.3) | 0.52 |
| dsDNA** | 127 (63.5) | 49 (46.7) | 69 (72.6) | <0.001** |
| aCL antibodies Freq (% positive) | 34 (17) | 10 (9.5) | 24 (25.3) | 0.003** |

\* $P<0.05$ ; \*\* $P<0.001$ ; Pearson's Chi-squared $P$ values are reported. For variables with expected cell counts <5, Fischer's Exact $P$ values are reported. For continuous variables (age and duration of follow-up) t-test $P$ values are reported. ESR=Erythrocyte sedimentation rate; CRP= C-Reactive Protein; ANA= Anti-nuclear antibodies; aCL= Anti-cardiolipin antibodies (IgG and/or IgM). Gastrointestinal involvement includes pancreatitis and hepatitis.

Most frequent antibodies were anti-dsDNA (63%), SSA (24%) and Ku (17.5%). Most common SLE clinical features in this population were arthritis (n=169, 85%), rash (n=111, 35%), alopecia (n=107, 53%) and cytopenias (n=89, 44.5%). Interstitial lung disease (ILD) was present in 10% (n=20) of our cohort. Neurological disease (ND, including brain lesions, transverse myelitis and mononeuritis multiplex) was present in 17.5% (n=35) and lupus nephritis was found in 16.5% (n=33) patients. Lupus nephritis, ND, oral ulcers, cytopenias, and gastrointestinal (GI) involvement (pancreatitis and hepatitis) were associated with severe SLE (Table 1). ANA speckled pattern was associated with mild to moderate disease (P=0.02). aCL (P<0.001) and dsDNA antibodies (P=0.003) were associated with severe disease. Elevated ESR (>30 mm/hour) and CRP (>0.5mg/dL) were more frequent in severe SLE. Median ESR was 31.5, whereas the maximum recorded ESR was 135 mm/hour. The 75^th^ percentile ESR was 60 mm/hour in our cohort. Median CRP was 0.8 mg/dL while the maximum was 29.3mg/dL.

From the univariate analysis, 15 predictors of severity were included in a multivariable stepwise cumulative logit model. The final model had 6 variables, namely gender, oral ulcers, alopecia, nephritis, ESR and aCL antibodies (Table 2). The odds of severe disease in males is about four times that of females (95% CI: 1.2, 13.5). There was a 4.5-fold increase in nephritis in severe SLE (95% CI: 1.9, 11.4). Patients with severe disease were 7 times more likely to suffer oral ulcers (95% CI: 2.8, 17.1) as compared to patients with mild to moderate SLE. The severe SLE group had significantly higher odds of presence of aCL antibodies (OR 2.4, 95% CI 1.0, 5.9). There were no interaction effects among the variables. The final model fit well (HL χ^2^ =9.8 *P*=0.2) and was as follows:

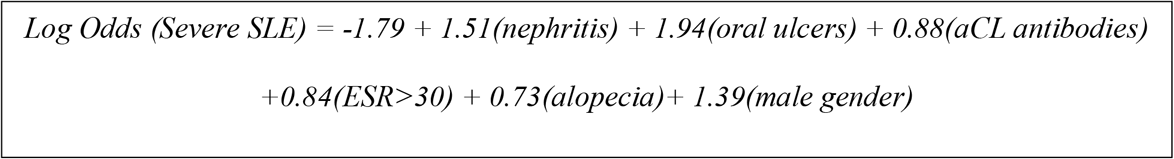

**Table 2:** Predictors of severe SLE.

| **Variable** | **Reference Level** | **Crude OR** | **95% CI** | **P value** | **Adjusted OR** | **95% CI** | **P value** |
| --- | --- | --- | --- | --- | --- | --- | --- |
| Oral Ulcers | Absent | 5.9 | (2.5, 13.6) | 0.001 | 6.9 | (2.8, 17.1) | 0.01* |
| Nephritis | Absent | 3.6 | (1.6, 8.2) | 0.002 | 4.5 | (1.9, 11.4) | 0.002* |
| Gender | Female | 1.9 | (0.7, 5.6) | 0.2 | 4.1 | (1.2, 13.5) | 0.02* |
| aCL IgM / IgG | Absent | 3.2 | (1.4, 7.1) | 0.004 | 2.4 | (1.0, 5.9) | 0.05* |
| ESR (mm/hour) | <30 | 2.6 | (1.4, 4.5) | 0.001 | 2.3 | (1.2, 4.4) | 0.01* |
| Alopecia | Absent | 1.5 | (0.9, 2.7) | 0.1 | 2.1 | (1.0, 4.1) | 0.04* |
| Cytopenias | Absent | 2.4 | (1.4, 4.3) | 0.002 | 1.6 | (0.8, 3.1) | 0.2 |
| CRP (mg/dL) | <0.5 | 2.2 | (1.3, 4.1) | 0.006 | 1.5 | (0.8, 3.0) | 0.2 |
| Subsets Factor 3 | Absent | 1.7 | (1.1, 2.6) | 0.02 | 1.6 | (1.0, 2.6) | 0.07 |
| Subsets Factor 2 | Absent | 1.3 | (0.9, 1.8) | 0.2 | 1.1 | (0.7, 1.6) | 0.7 |
| GI | Absent | 2.4 | (1.6, 8.2) | 0.05 | 1.8 | (0.6, 5.2) | 0.3 |
| Rash | Absent | 1.4 | (0.8, 2.4) | 0.2 | 1.4 | (0.7, 2.7) | 0.3 |
| Clots | Absent | 2.3 | (0.6, 9.4) | 0.25 | 1.3 | (0.2, 6.6) | 0.8 |
| ANA-H | Absent | 1.5 | (0.9, 2.6) | 0.15 | 0.9 | (0.5-1.9) | 0.9 |
| ANA-speckled | Absent | 0.3 | (0.1, 0.8) | 0.02 | 0.4 | (0.2, 1.3) | 0.2 |

\*P value <0.05 at the multivariable level of analysis. ANA-H: Homogenous pattern; Subsets Factor 2: SSA, SSB; Subsets Factor 3: dsDNA; GI: Gastrointestinal involvement includes pancreatitis and hepatitis.

Overall, clinical remission was achieved in 62% of the total cohort of 200 patients. However, remission outcomes were formally assessed in 94 patients (47%) followed for over a year. Clinical remission was achieved in 82 patients (87%), out of which 37 were in severe SLEDAI category. The mean time to remission is 28.9±21.3 months. The mean duration of follow-up in this subset is 41±19 months. For every one month increase in duration of follow-up, the hazard of time to remission increases by 4% (95% CI 0.95-0.98; *P*<0.001). The Kaplan Meier survival curve (Figure 2) demonstrated that the hazard of time to remission is 48% less in severe SLE as compared to mild-moderate disease (95% CI 0.33-0.82; *P*=0.005).

**Figure 2:**
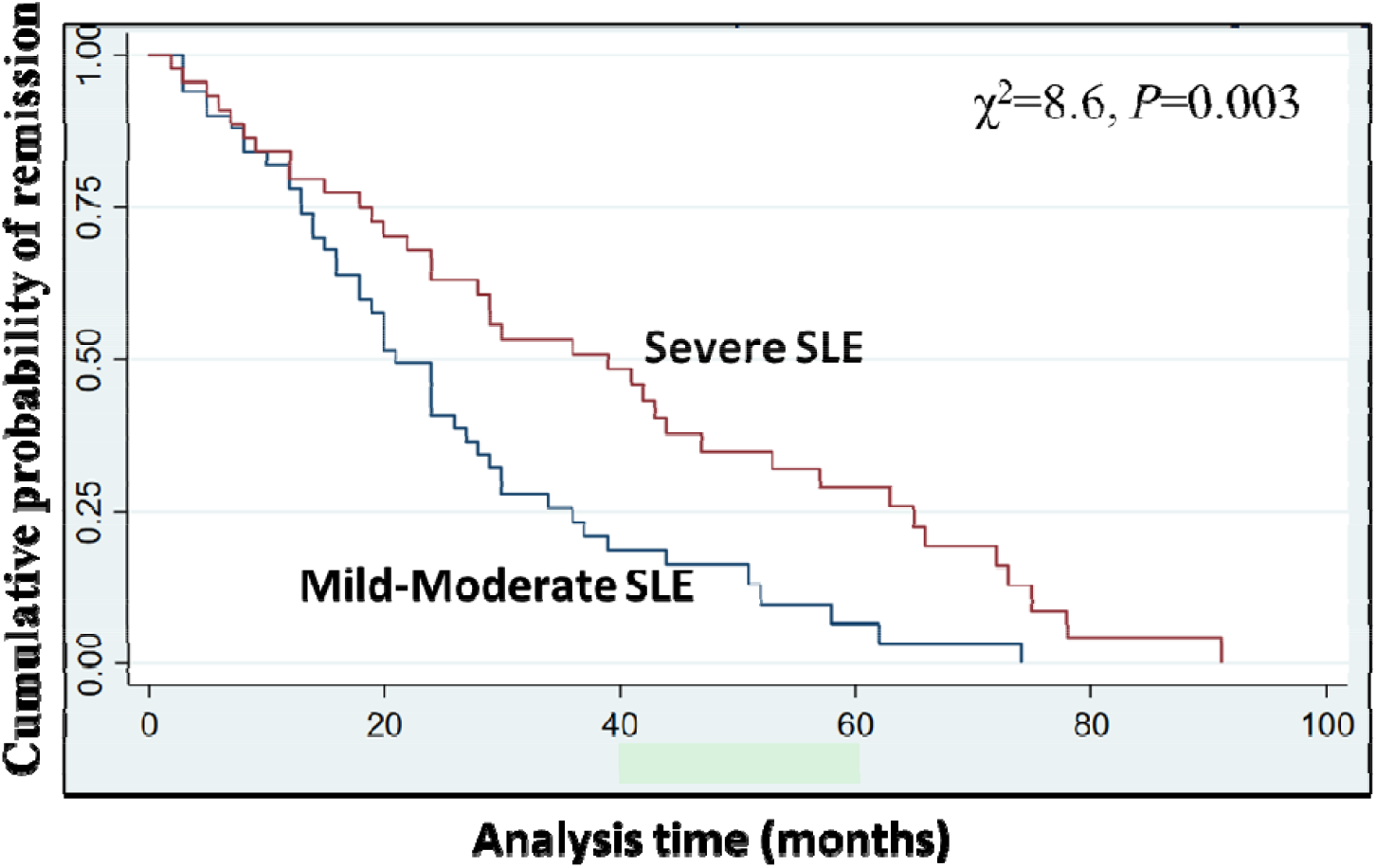
Kaplan Meier curve for remission time in SLE patients followed for >1-year (n=94). Log rank test was used to test for equality of the two survivor functions.

As shown in Table 3, 21% (n=20) patients (12 originally in severe category) decided to discontinue their immunosuppression completely after being in clinical remission for over 2-years (ROFT). ROFT was achieved in 27.3±13.1 months, with a maximum of 54 months. All of them were on daily Pred <2.5mg and HCQ 200mg. No flare was reported for a mean post-remission follow-up of 18±15 months. Patients who achieved remission required higher mean doses of Pred, while requiring lower mean doses of azathioprine (AZA), mycophenolate (MMF) and Rituximab (RTX). Overall median daily dose of Pred was 5.7mg while 75^th^ percentile was 9.3mg, indicating that majority of the patients were under 10mg daily dose of Pred. Higher Pred equivalent doses were mainly due to methylprednisolone pulses. Mean duration of Pred use is 21.4 months. In 2 patients (in RONT group), HCQ was discontinued due to GI intolerance.

**Table 3:** Doses of Immunosuppressants by remission pattern.

| **Medication** | **Remission not achieved**<br>**n=12** | **RONT**<br>**n=62** | **ROFT**<br>**n=20** |
| --- | --- | --- | --- |
| Pred (mg/day) | 12.7 ± 8.3 | 16 ± 28.3 | 16 ± 31.6 |
| HCQ (mg/day) | 283.0 ± 70.4 | 308.6 ± 64.5 | 294.5 ± 74.2 |
| AZA (mg/day) | 175.8 ± 222.2 | 99.9 ± 44.7 | 85.7 ± 23.0 |
| CsA (mg/day) | 50.0 ± 0 | 49.7 ± 10.7 | 76.7 ± 36 |
| MTX (mg/week) | 10.4 ± 6.3 | 43.5 ± 89.9 | 27.0 ± 18.7 |
| MMF (g/day) | 1.35 ± 0.38 | 1.18 ± 0.35 | 1.14 ± 0.43 |
| Cyc (total dose g) | 3.25 ± 2.57 | 2.74 ± 1.55 | 4.25 ± 0 |
| RTX (total dose g) | 2.5 ± 2.0 | 1.73 ± 0.87 | 1.86 ± 0.95 |

Medication doses: Mean ± standard deviation. Pred: Prednisolone. HCQ: Hydroxychloroquine. AZA: Azathioprine. CsA: Ciclosporin A. MTX: Methotrexate. MMF: Mycophenolate mofetil. RTX: Cyc: Cyclophosphamide. Rituximab. RONT: Remission on treatment. ROFT: Remission off treatment.

Factor analysis reduced clinical presentations of SLE to 4 major subtypes, *viz* ND, nephritis, cytopenias and alopecia (Figure 3a). These four factors together explained 34.4% of the variance in clinical presentations in our SLE patients. Factor analysis reduced SLE associated antibodies to 4 major factors. First, Mi2, Ku, RNP, NUC, Histones, AMA-M2 (41.6% variance explained), second SSA, SSB (34.2% variance explained) and third, the dsDNA antibody (22.8% variance explained) and fourth being absence of subset antibodies. These 4 factors together explained 23.6% of the variance in antibody associations in our SLE cohort (Figure 3b).

**Figure 3.**
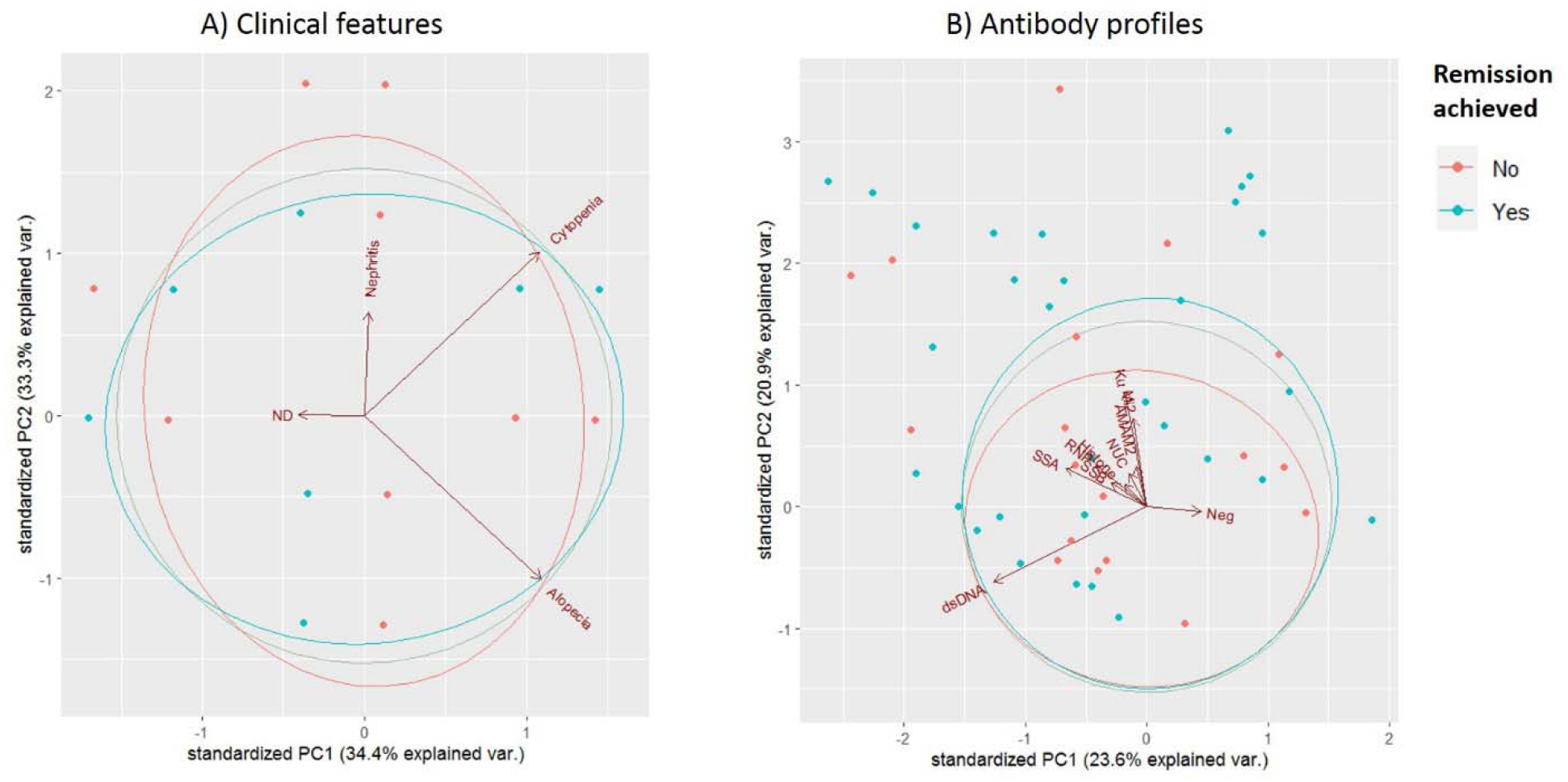
Factor analysis of SLE A) Clinical features B) Antibodies. 3A) Factor 1: ND (Neurological disease), Factor 2: Nephritis, Factor 3: Cytopenia (anemia, leucopenia or thrombocytopenia), Factor 4: Alopecia. 3B) Factor 1: Mi2, Ku, RNP, Histones, NUC: Nucleosome antibodies and AMAM2: Anti-mitochondrial antibody M2. Factor 2: SSA, SSB antibodies. Factor 3: dsDNA antibody. Factor 4: Neg: no ANA subsets antibodies detected.

## Discussion

Systemic Lupus Erythematosus (SLE) is a difficult to treat autoimmune disease due to clinical heterogeneity, unpredictability of disease flares and limited therapeutics. These challenges are worsened in LMIC. This study is an analysis of real-world data in an LMIC setting and presents a uniquely detailed description of SLE disease patterns and remission in Pakistan. Of the 200 complete patient records analyzed, 47.5% suffered from severe SLE. Since ImmunoCure is a Rheumatology referral center, the high percentage of severe SLE may be due to referral bias. Age and gender distribution was comparable to worldwide studies (5, 9, 30, 31). ILD was present in 10%, ND in 17.5% and nephritis in 16.5% of our cohort. Previously, the estimated frequency of ILD in SLE patients has been reported to be between 3% and 9% (32), and ILD has been shown to be associated with increasing age and disease severity (33).

Though dsDNA was the commonest antibody, there was substantial positivity for anti-Ku antibody with a trend towards association with mild-moderate SLE (*P*=0.09). Interestingly ANA speckled pattern was associated with mild-moderate SLE. Usually, speckled pattern is generated by the anti-Sm antibody (34) that is associated with severe SLE (35). However, none of the antibodies were associated with severe SLE in our cohort except aCL (OR 2.4; 95% CI 1.0-5.9). Recently, aCL antibodies were shown to be associated with severe SLE in a Chinese cohort (17) and we confirm this finding here.

Initial ESR>30 and CRP>0.8mg/dL were indicative of severe SLE. These are easily measurable laboratory parameters that can alert the physician to the possibility of severe SLE early in the course of the disease. Severe SLE was strongly associated with clinical presentation of oral ulcers, alopecia, nephritis and aCL antibodies and ESR>30mm/hour. High CRP did not make it into this multivariate analysis model. Oral ulcers and alopecia are easily observable clinical findings that have been shown here to associate strongly with severe disease. Hence, with the help of a basic clinical exam together with a simple blood test (ESR) and a urine detailed report, severe SLE can be identified early, even in a general practice clinic setting. Since arthritis was present in 85% of our patients, complaints of joint pains and morning stiffness in the general population would assume special importance in possible SLE diagnosis. Male gender and nephritis are known to be strongly associated with severe SLE (36). Hence, we calculated the predicted odds of severe SLE for the common scenario of a female presenting with oral ulcers, alopecia and ESR>30mm/hour. Using our model, we found the predicted odds for severe SLE in such a case to be 5.6. This kind of clinical approach is critical as SLE often presents vaguely and diagnosis is delayed, leading to complications associated with increased mortality from nephritis, increased risk of cardiovascular disease and infections (3). Moreover, in a LMIC setting these findings are cost-effective to identify.

Most SLE patients do achieve remission (37). Previously reported remission rates range between 35% and 88% (24, 37, 38). In a Chinese study of 218 SLE patients RONT was reported at 67.9% and ROFT 10.6% whereas in an Italian study (n=155), RONT was reported to decrease over the course of 5-years from 45% to 22% and ROFT from 12% initially to 5.2% (19, 39). We report RONT in 66% and ROFT in 21% 94 SLE patients followed for over a year. In Pakistan, as with many traditional cultures, family and friend support are an integral part of the social framework, and this may have contributed significantly to the high remission rates reported here. Earlier studies have reported lack of social support to be associated with poor outcomes in SLE (7). At ImmunoCure, the patient is nearly always accompanied to the clinic by family or friends. Both remission status and follow-up duration are associated with the social support systems. These studies, together with ours, build hope that remission in SLE is substantially frequent and even ROFT may be possible given current treatment regimens.

Long term steroid therapy is a dilemma in SLE therapy (40). In our study those who achieved remission, received higher total doses of Pred mainly due to prolonged gradual taper. Pred withdrawal after RONT is debated and a recent randomized clinical trial concluded against this strategy noting significantly increased flares (40). Steroid withdrawal assumes critical importance in LMIC, where patients face multiple barriers to healthcare, including access to Rheumatologists, availability of biologics and financial difficulties. In Pakistan, patients travel long distances (9-to-12-hour journey) to major cities like Karachi for Rheumatology care. Prior to COVID19 telemedicine was not frequently used in Rheumatology and still has risks over in-clinic consultations (41). However, with the help of telemedicine routine follow-up can be improved to a degree and may lead to higher remission rates. Belimumab, anifrolumab and tofacitinib were not available in Pakistan during the period of this study. Their addition along with newer SLE drugs in the clinical pipeline will likely enhance the possibility of early and prolonged clinical remission and minimization of steroid use (42, 43).

Based on ACR 1997 criteria, 330 types of SLE are possible (44). We attempted to minimize this heterogeneity by evaluating SLE subgroups of clinical and antibody profiles in our population. Reduction of clinical heterogeneity in SLE will likely help streamline treatment strategies as well. We show using factor analysis that SLE in our cohort demonstrated 4 clinical and 3 antibody-based subtypes. Recently, Diaz-Gallo et al. showed that 4 SLE subgroups based on autoantibody profile could be distinguished (13). Idborg et al. showed that there were differences at the proteomic level based on SLE antibody profile (15). Therefore, stratifying SLE patients based on clinical and antibody profiles may help to better understand the pathobiology of SLE as well as improve targeted therapy for the disease.

The strengths of this study lie in its longitudinal design and detailed description of SLE from Pakistan. The results of our study are widely applicable to LMICs, and to SLE long term care and remission worldwide. Moreover, we describe subtypes of SLE that help to reduce clinical heterogeneity of SLE and present a clinical model for rapidly determining severe disease in the clinic. There were some limitations in this study. First, this was a single-center study with relatively moderate sample size. Second, accrual of damage and flare rate was not evaluated.

Third, no predefined protocol for de-escalation of therapy is described but was clinically guided. The expansion of the study in terms of sample size and follow-up duration, with data on flares is warranted. Our data pave the way towards T2T remission strategies in patients with SLE in LMICs, reinforcing their relevance in clinical practice.

## Data Availability

All data produced in the present work are contained in the manuscript

